# Zero-shot Interpretable Phenotyping of Postpartum Hemorrhage Using Large Language Models

**DOI:** 10.1101/2023.05.31.23290753

**Authors:** Emily Alsentzer, Matthew J Rasmussen, Romy Fontoura, Alexis L Cull, Brett Beaulieu-Jones, Kathryn J Gray, David W Bates, Vesela P Kovacheva

**Affiliations:** Division of General Internal Medicine and Primary Care, Brigham and Women’s Hospital, Boston, USA; Department of Anesthesiology, Perioperative and Pain Medicine, Brigham and Women’s Hospital, Boston, MA; Section of Biomedical Data Science, Department of Medicine, University of Chicago, Chicago, IL; Centerfor Genomic Medicine, Massachusetts General Hospital, Boston, MA; Division of Maternal-Fetal Medicine, Brigham and Women’s Hospital, Boston, MA; Department of Health Care Policy and Management, Harvard T. H. Chan School of Public Health, Boston, USA

## Abstract

Many areas of medicine would benefit from deeper, more accurate phenotyping, but there are limited approaches for phenotyping using clinical notes without substantial annotated data. Large language models (LLMs) have demonstrated immense potential to adapt to novel tasks with no additional training by specifying task-specific i nstructions. We investigated the performance of a publicly available LLM, Flan-T5, in phenotyping patients with postpartum hemorrhage (PPH) using discharge notes from electronic health records (*n*=271,081). The language model achieved strong performance in extracting 24 granular concepts associated with PPH. Identifying these granular concepts accurately allowed the development of interpretable, complex phenotypes and subtypes. The Flan-T5 model achieved high fidelity in phenotyping PPH (positive predictive value of 0.95), identifying 47% more patients with this complication compared to the current standard of using claims codes. This LLM pipeline can be used reliably for subtyping PPH and outperformed a claims-based approach on the three most common PPH subtypes associated with uterine atony, abnormal placentation, and obstetric trauma. The advantage of this approach to subtyping is its interpretability, as each concept contributing to the subtype determination can be evaluated. Moreover, as definitions may change over time due to new guidelines, using granular concepts to create complex phenotypes enables prompt and efficient updating of the algorithm. Using this language modelling approach enables rapid phenotyping without the need for any manually annotated training data across multiple clinical use cases.

---

Robust phenotyping is essential to multiple clinical and research workflows, such as clinical diagnosis [1], novel phenotype discovery [2], clinical trial screening [3], comparative effectiveness research [4], quality improvement [5], and genome-wide and phenome-wide association studies [6]. The wide adoption of electronic health records (EHR) has enabled the development of digital phenotyping approaches, which seek to leverage the large amounts of electronic patient data, stored as structured data (*e.g.* diagnosis codes, medications, and laboratory results) and unstructured clinical notes, to characterize a patient’s clinical presentation. Currently, many phenotyping approaches utilize diagnosis codes such as the International Classification of Diseases (ICD) [7] or a combination of rules based on structured data [8, 9]. While structured data is readily available and easily computable, structured data often reflect billing processes rather than disease course and do not capture the nuanced clinical narrative found in the EHR notes [10, 11].

Natural language processing (NLP) methods have increasingly been used to enable more precise, multi-modal phenotyping by automating data extraction from unstructured clinical notes. Most approaches are rule-based, relying on keywords [12], regular expressions [13], and/or existing NLP tools that extract medical ontology concepts [14,15] to identify relevant information from notes. These approaches produce interpretable phenotypes, but often have low recall due to the use of hand-crafted features. In contrast, supervised machine learning based phenotyping approaches allow for learning useful features and may produce less brittle models [11,16,17]. However, supervised machine learning models have been difficult to implement because they require substantial clinician-annotated data for training robust models.

Recent advances in training large language models (LLMs) offer an opportunity to develop generalizable phenotypes without substantial annotated data. A common paradigm in NLP has been to first pretrain a model via a self-supervised learning objective on large amounts of unlabeled text and then fine-tune the model on a specific downstream task of interest using labeled training data. However, more recent work has demonstrated the ability of LLMs to adapt to new tasks without any gradient updates or fine-tuning, simply by specifying task instructions via text interaction with the model [18–20]. For example, Agrawal et al. found that GPT-3 can perform few-shot information extraction on several clinical tasks, including acronym disambiguation, co-reference resolution, and medication extraction [21]. Contemporaneous to this work, McInerney et al. show that Flan-T5 can perform feature extraction from clinical notes to enable training of risk prediction models [22]. The zero- and few-shot capabilities of these models (*i.e.*, their ability to generalize to novel tasks with zero or limited examples) can enable rapid development of models for diverse clinical applications where labeled data is sparse or expensive to acquire.

In this work, we investigate the utility of LLMs for zero-shot phenotyping using clinical notes. As a proof of concept, we focus on phenotyping postpartum hemorrhage (PPH), the leading cause of severe maternal morbidity and mortality. Globally, PPH complicates 2-3% of all pregnancies and accounts for 140,000 maternal deaths annually [23]. The current PPH definition is based on the documentation of estimated blood loss of at least 1000 mL following delivery [24]. PPH is typically identified in large epidemiological studies using diagnosis codes [7, 25]; however, such codes have limited accuracy [7]. More comprehensive PPH phenotype definitions have been developed [8], but there is limited work in incorporating clinical notes into PPH phenotype definitions [26]. Furthermore, several subtypes of PPH exist, each of which is associated with different etiologies, management, and outcomes. Clinical guidelines recommend prompt identification of the PPH subtype of PPH in order to initiate appropriate interventions [24]. However, to the best of our knowledge, there are no digital phenotyping algorithms for PPH subtypes.

The goal of this study was to develop a robust and interpretable approach for rapid phenotyping and subtyping of postpartum hemorrhage (PPH) patients. We leveraged Flan-T5, an open-source LLM, to perform zero-shot extraction of PPH-related concepts from obstetric discharge summaries, and we used the extracted concepts to perform interpretable PPH phenotyping and subtyping. We validated the extracted concepts using manual expert chart review and found that Flan-T5 can extract PPH-related concepts with strong performance and can identify PPH patients and subtypes of PPH that are missed by common structured data approaches. This two-step extract-then-phenotype approach allows for greater interpretability compared to end-to-end supervised machine learning models and better recall compared to more brittle keyword or regex rules-based models. Furthermore, the open-source Flan-T5 LLMs can be run locally behind a hospital firewall, limiting concerns regarding the security, privacy, and run-time costs of the models. Our findings highlight the potential for LLMs to annotate clinical notes and facilitate rapid and accurate phenotyping for diverse clinical outcomes.

## Results

### Study cohort

We identified 138,648 unique individuals with an obstetric encounter resulting in birth after 20 gestational weeks at the Mass General Brigham (MGB) hospitals in Boston, MA, from 1998-2015. There were a total of 503,991 discharge summaries for patients in this initial cohort. We leveraged discharge summaries for NLP-based phenotyping, as postpartum hemorrhage (PPH) typically develops within 24 hours after delivery, and patients are not discharged until at least 24 hours post-delivery. As not every discharge note described a delivery encounter, we developed a combination of terms to identify delivery-related discharge notes with high recall (See Methods). After filtering discharge summaries that did not describe a delivery encounter, our final study cohort was comprised of 131,284 unique patients with 271,081 discharge summaries (Figure 1a). Each patient had an average of 2.1 obstetric discharge summary notes (SD = 1.4), consisting of 807.6 words on average (SD = 923.1). The cohort characteristics are summarized in Figure 1b-e.

**Figure 1:**
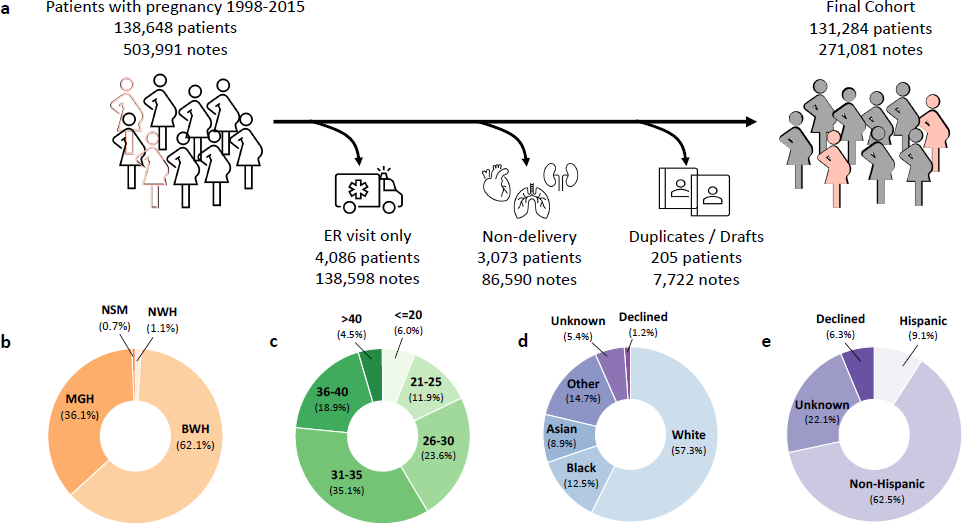
Obstetric delivery cohort. **(a)** Inclusion criteria for identifying delivery-related discharge summaries. The final cohort consists of 271,081 discharge notes for 131,284 women with an obstetric encounter at Mass General Brigham hospitals from 1998-2015. **(b-e)** Characteristics of the cohort based on **(b)** delivery hospital and **(c)** patient age at delivery, **(d)** patient race, and **(e)** patient ethnicity. ER, emergency room; MGH, Massachusetts General Hospital; BWH, Brigham and Women’s Hospital, NSM, North Shore Medical Center, NWH, Newton Wellesley Hospital

### Zero-shot extraction of PPH-related concepts with NLP

We developed a list of 24 PPH-related concepts based on an extensive literature review and discussion with expert physicians (V.K. and K.G.; full list and definitions in Supplemental Table 1). We identified the PPH-related concepts in all discharge summaries by prompting the 11 billion parameter Flan-T5 model (Flan-T5-XXL), to either answer a yes/no question regarding the presence of a condition, medication, or procedure (*e.g.*, uterine atony) or to extract a specific measurement (*e.g.*, estimated blood loss; Figure 2a). This approach allows for extraction of concepts from unstructured text without requiring data to further train the model.

**Figure 2:**
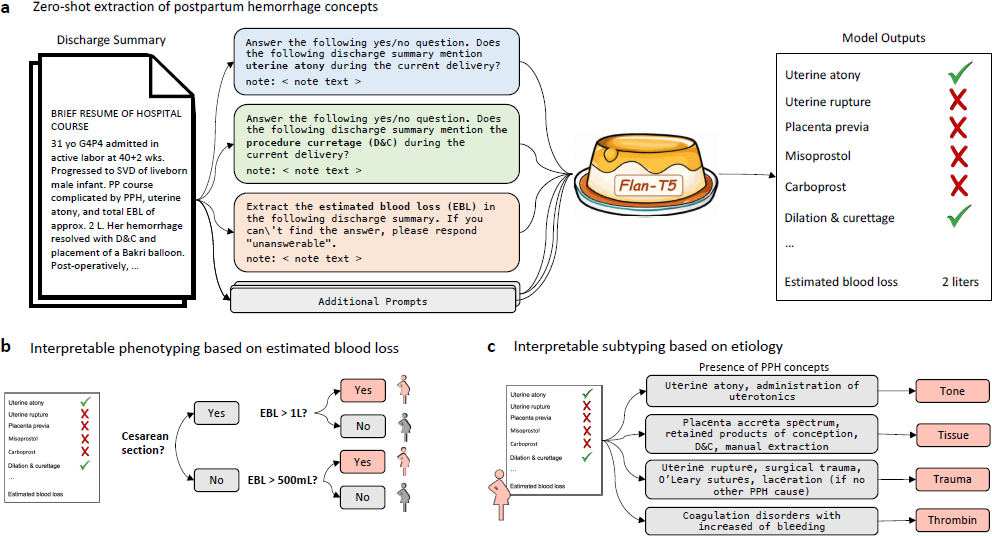
Overview of zero-shot NLP pipeline for accurate and interpretable postpartum hemorrhage (PPH) pheno-typing. **(a)** Zero-shot extraction of PPH concepts using Flan-T5. We constructed either yes/no or extraction prompts for each PPH concept, concatenated the discharge summary of interest, and fed the combined input into Flan-T5. This process enabled the rapid extraction of PPH-related information from notes with no training data. **(b)** We leveraged the extracted concepts to perform interpretable phenotyping of PPH defined as cesarean deliveries with at least 1L or vaginal deliveries with at least 500mL of estimated blood loss. **(c)** We used the extracted concepts to perform interpretable subtyping of PPH based on the underlying etiology. We assigned a delivery note to the “tone”, “tissue”, “trauma”, or “thrombin” subtype if any of the concepts associated with the subtype are present. PPH, postpartum hemorrhage; EBL, estimated blood loss

We evaluated the Flan-T5 model performance on a random sample of 1,175 manually annotated discharge summaries with PPH ICD diagnosis codes. Discharge summaries with PPH diagnosis codes were used to enrich for obstetric discharge summaries more likely to contain PPH-related concepts. The Flan-T5 performance is summarized in Table 1. We compared the Flan-T5 NLP models to regular expressions constructed for each PPH-related concept (Figure 3; Supplemental Table 2). Regular expressions were selected as the baseline approach because they can similarly be constructed with minimal “training” data. Our primary evaluation metric for the classification tasks was binary F1 score, which is useful for imbalanced datasets with rare labels where performance on the positive class is most important. The Flan-T5 model achieved a binary F1 score of 0.75 or higher on 21 of 23 binary concepts, exceeding a 0.9 binary F1 score on 12 of the PPH concepts. Flan-T5 significantly outperformed the corresponding regular expression for nine binary PPH-related concepts (all p-values *<* 0.05). Furthermore, the model extracted estimated blood loss values with a recall of 0.745, precision of 0.905, and note level accuracy of 95.1%. While regular expressions achieved similar performance on simpler extraction tasks, such as identification of medication use, the Flan-T5 model outperformed regular expressions on concepts that are expressed in clinical notes in variable formats, such as coagulation disorders. The model was able to infer the presence of some concepts; however, this contributed to higher false positive rates if there was no explicit presence of the concept in the notes. For example, the notes that contained postpartum ‘dilation and curettage’ were also often predicted to be positive for ‘manual removal of placenta’; the latter procedure commonly precedes curettage. The false negatives were commonly due to unusual abbreviations or concepts with multiple misspellings. For example, in one of the notes containing the following text ‘500ccf/b d&c for retained POC’, the model correctly identified the concept ‘retained products of conception’ but was unable to extract the blood loss, in this case ‘500’ or identify ‘dilation and curettage’.

**Figure 3:**
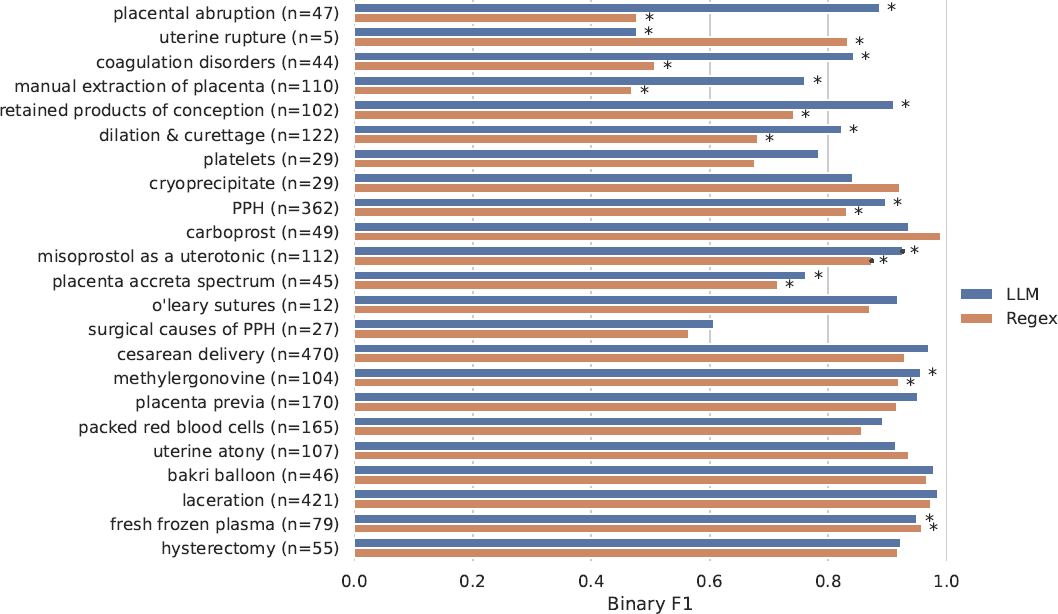
Comparison of the Flan-T5 language model and regular expression approaches for zero-shot extraction of postpartum hemorrhage (PPH)-related concepts. The prevalence of each concept in the annotated test set is reported and compared to the model performance according to binary F1 score. The stars (*) denote that there is a significant difference using the McNemar test (*p <* 0.05) between the regex and language model performance. PPH, postpartum hemorrhage.

**Table 1:** Performance of the zero-shot Flan-T5 model in identifying postpartum hemorrhage (PPH) concepts in dis-charge summaries. We assessed model performance on 1175 manually annotated notes with PPH ICD diagnostic codes. We report the prevalence of each concept in the annotated test set. We report model sensitivity, specificity, positive predictive value (PPV), binary F1 score, and accuracy for binary concepts, and we report sensitivity, PPV, and note-level accuracy in extracting the correct estimated blood loss values. PPH, postpartum hemorrhage

| Concept (n) | Sensitivity | Specificity | PPV | Binary F1 | Accuracy |
| --- | --- | --- | --- | --- | --- |
| Laceration (n=421) | 0.976 | 0.996 | 0.993 | 0.984 | 0.989 |
| Bakri balloon (n=46) | 0.957 | 1.000 | 1.000 | 0.978 | 0.998 |
| Cesarean delivery (n=470) | 0.983 | 0.970 | 0.957 | 0.970 | 0.975 |
| Methylergonovine (n=104) | 0.952 | 0.996 | 0.961 | 0.957 | 0.992 |
| Placenta previa (n=170) | 0.982 | 0.986 | 0.923 | 0.952 | 0.986 |
| Fresh frozen plasma (n=79) | 0.962 | 0.995 | 0.938 | 0.950 | 0.993 |
| Carboprost (n=49) | 0.898 | 0.999 | 0.978 | 0.936 | 0.995 |
| Misoprostol as a uterotonic (n=112) | 0.938 | 0.991 | 0.913 | 0.925 | 0.986 |
| Hysterectomy (n=55) | 0.982 | 0.993 | 0.871 | 0.923 | 0.992 |
| O'Leary sutures (n=12) | 0.917 | 0.999 | 0.917 | 0.917 | 0.998 |
| Uterine atony (n=107) | 0.850 | 0.999 | 0.989 | 0.915 | 0.986 |
| Retained products of conception (n=102) | 0.892 | 0.993 | 0.929 | 0.910 | 0.985 |
| Direct mention of PPH (n=362) | 0.881 | 0.963 | 0.914 | 0.897 | 0.938 |
| Packed red blood cells (n=165) | 0.945 | 0.971 | 0.843 | 0.891 | 0.968 |
| Abruption of the placenta (n=47) | 0.915 | 0.994 | 0.860 | 0.887 | 0.991 |
| Coagulation disorders (n=44) | 0.795 | 0.996 | 0.897 | 0.843 | 0.989 |
| Cryoprecipitate (n=29) | 1.000 | 0.990 | 0.725 | 0.841 | 0.991 |
| Dilation and curettage (n=122) | 0.762 | 0.990 | 0.894 | 0.823 | 0.966 |
| Platelets (n=29) | 1.000 | 0.986 | 0.644 | 0.784 | 0.986 |
| Placenta accreta spectrum (n=45) | 0.822 | 0.987 | 0.712 | 0.763 | 0.980 |
| Manual extraction of placenta (n=110) | 0.782 | 0.972 | 0.741 | 0.761 | 0.954 |
| PPH due to surgical causes (n=27) | 0.741 | 0.983 | 0.513 | 0.606 | 0.978 |
| Uterine rupture (n=5) | 1.000 | 0.992 | 0.357 | 0.526 | 0.992 |
| Concept (n) | Sensitivity | PPV | Note Acc. |  |  |
| Estimated blood loss (n=212) | 0.745 | 0.905 | 0.951 |  |  |

While prompts were developed solely on notes from a single hospital site, we found that the model generalized well to notes from other MGB hospitals. There was no substantial difference in performance across hospital sites (Supplemental Table 3).

### NLP-based Identification of PPH cases

We leveraged the extracted concepts to identify deliveries with postpartum hemorrhage (Figure 2b). The Flan-T5 model extracted the estimated blood loss values and delivery type (cesarean or vaginal) from all notes in the cohort (*n* = 271,081). Subsequently, we classified notes as describing PPH if patients had blood loss greater than 500 mL for vaginal deliveries or greater than 1000 mL for cesarean deliveries. This matched the PPH definition used in the hospitals during the study period (See Methods). There were 2,598 discharge summary notes for 2,270 deliveries in our study cohort with PPH, according to the model. The prevalence of each PPH-related concept in the deliveries with PPH identified based on model-extracted blood loss can be found in Table 2.

**Table 2:** Prevalence of each postpartum hemorrhage (PPH)-related concept in the deliveries with predicted PPH. Both the count and percentage of each concept are reported. The prevalence is reported in the 2,270 deliveries that the natural language processing (NLP)-based model labelled as describing postpartum hemorrhage. PPH, postpartum hemorrhage;

| Concept | Prevalence (%) |
| --- | --- |
| Cesarean delivery | 1002 (44.14) |
| Direct mention of PPH | 915 (40.31) |
| Laceration | 735 (32.38) |
| Packed red blood cells | 681 (30.00) |
| Manual extraction of placenta | 532 (23.44) |
| Misoprostol as a uterotonic | 361 (15.90) |
| Dilation and curettage | 339 (14.93) |
| Methylergonovine | 325 (14.32) |
| Fresh frozen plasma | 319 (14.05) |
| Retained products of conception | 302 (13.30) |
| Uterine atony | 261 (11.50) |
| Placenta previa | 201 (8.85) |
| Hysterectomy | 185 (8.15) |
| Carboprost | 159 (7.00) |
| Platelets | 147 (6.48) |
| Placenta accreta spectrum | 141 (6.21) |
| PPH due to surgical causes | 135 (5.95) |
| Bakri balloon | 132 (5.82) |
| Coagulation disorders | 129 (5.68) |
| Abruption of the placenta | 129 (5.68) |
| Cryoprecipitate | 113 (4.98) |
| Uterine rupture | 106 (4.67) |
| O'Leary sutures | 40 (1.76) |

We evaluated the PPH phenotyping algorithm by comparing the model performance on a random sample of 300 expert-annotated discharge summary notes predicted by Flan-T5 to describe deliveries with PPH. The gold-standard discharge summaries were annotated via manual medical record review. The positive predictive value of the NLP-based PPH phenotyping algorithm is 0.95. The NLP-based approach allows identification of PPH cases that lack any PPH International Classification of Diseases (ICD) diagnosis codes associated with the delivery. Of the 285 discharge summaries with confirmed PPH, 148 and 136 did not have an associated PPH ICD code according to the definitions in Butwick et al. [7] and Zheutlin et al. [8], respectively. Over 47% of the discharge summaries with PPH would not have been identified using PPH ICD codes alone.

We further examined the relationship between model-extracted estimated blood loss and presence of PPH ICD codes on the entire notes cohort (*n* = 271,081). As the blood loss amount increases, there is more likely to be a PPH ICD code associated with the admission, as expected (Fig. 4, Supplemental Figure 2). When no estimated blood loss is extracted, the prevalence of PPH ICD codes is less than 5%. When the extracted blood loss values are between 1000 and 1500mL, criteria which meet both historical and current definitions of PPH, less than 50% of notes have an associated PPH ICD code.

**Figure 4:**
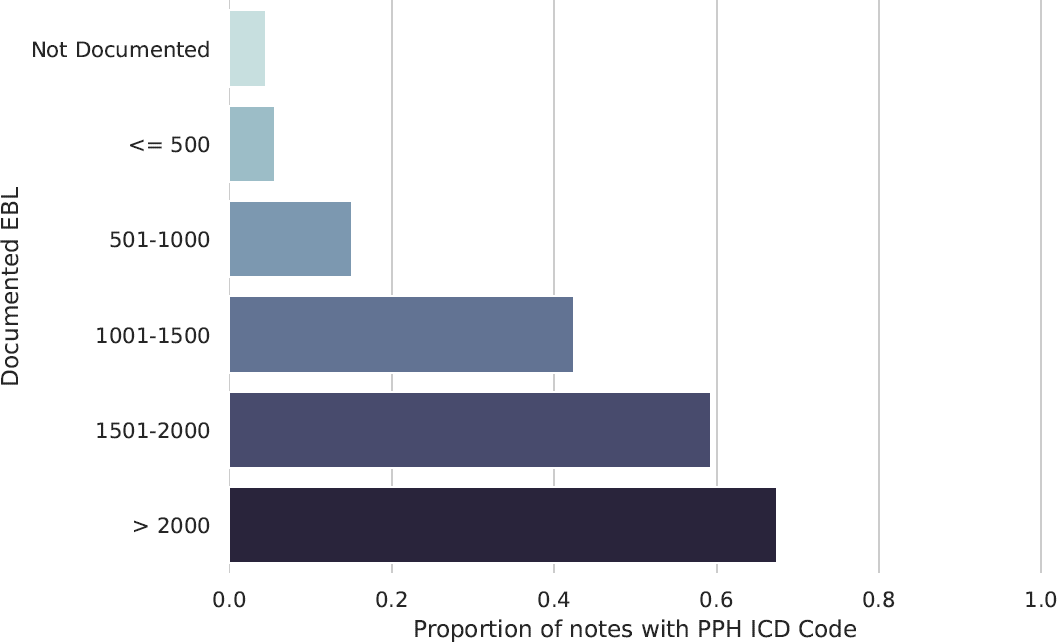
Prevalence of postpartum hemorrhage (PPH) international classification of diseases (ICD) codes in notes with varying estimated blood loss (EBL) The *y*-axis depicts the range of estimated blood loss (EBL) values extracted from the delivery note by Flan-T5 (if any), and the *x*-axis denotes the proportion of notes in each EBL category with a postpartum hemorrhage ICD diagnostic code. PPH ICD codes are defined according to the definition in Zheutlin et al. [8]. Refer to supplemental Figure 2 for a similar plot using the PPH ICD definition from Butwick et al. [7]. PPH, postpartum hemorrhage; ICD, international classification of diseases; EBL, estimated blood loss.

### NLP-based Classification of PPH subtypes

Finally, we leverage the extracted PPH concepts to classify deliveries with PPH into four subtypes corresponding to distinct causes of PPH: uterine atony (“tone”), retained products of conception and placenta accreta spectrum, (“tissue”), birth or surgical trauma, (‘trauma’) and coagulation abnormalities (“thrombin”; Figure 2E). We constructed composite phenotypes for each subtype based on the presence of one or more NLP-extracted PPH terms (See Methods). Using these NLP-based subtyping algorithms, we estimate that 29.9% of predicted PPH deliveries in our study cohort are due to uterine atony, 26.8% are due to retained products of conception, 24.0% are due to trauma, and 5.7% are due to coagulation abnormalities.

We validated the subtyping algorithm via an expert manual medical record review of the 285 confirmed PPH deliveries. The PPH subtype could be determined via manual review of the discharge summary in 73% of notes. We found that the NLP-based approach can achieve strong performance in subtype classification of those notes across three of the four subtypes, significantly outperforming an ICD-based approach to PPH subtyping for the “tone”, “tissue”, and ”trauma” subtypes with binary F1 score by 50.6%, 66.5%, and 17.8%, respectively (p-value *<* 0.01; Table 3).

**Table 3:**
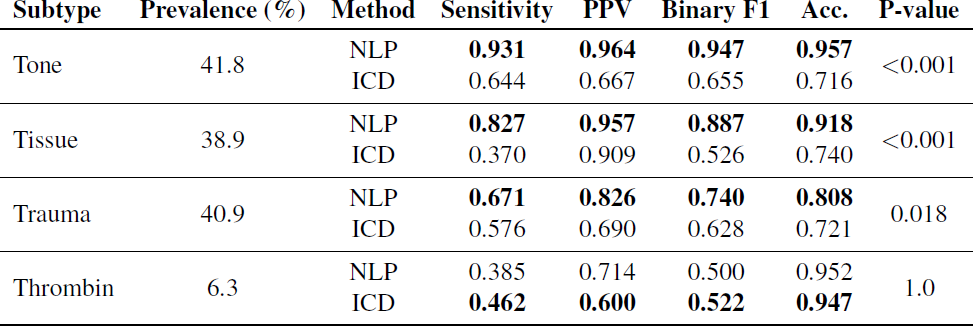
Comparison of NLP and ICD approaches for PPH subtyping. The true prevalence of each subtype and the NLP and ICD model performance are shown. Note that a single episode of postpartum hemorrhage can be classified with multiple subtypes. The model evaluation was performed on 285 notes with confirmed PPH via manual annotation. In 27.0% of the notes, the annotators were unable to determine a PPH subtype from the discharge summary alone (not shown). These notes were excluded to avoid inflation of false positives in the ICD model in cases where subtype information was captured elsewhere in the medical record. NLP, natural language processing; PPH, postpartum hemorrhage; ICD, international classi-fication of diseases.

| Subtype | Prevalence (%) | Method | Sensitivity | PPV | Binary F1 | Acc. | P-value |
| --- | --- | --- | --- | --- | --- | --- | --- |
| Tone | 41.8 | NLP | **0.931** | **0.964** | **0.947** | **0.957** | <0.001 |
|  |  | ICD | 0.644 | 0.667 | 0.655 | 0.716 |  |
| Tissue | 38.9 | NLP | **0.827** | **0.957** | **0.887** | **0.918** | <0.001 |
|  |  | ICD | 0.370 | 0.909 | 0.526 | 0.740 |  |
| Trauma | 40.9 | NLP | **0.671** | **0.826** | **0.740** | **0.808** | 0.018 |
|  |  | ICD | 0.576 | 0.690 | 0.628 | 0.721 |  |
| Thrombin | 6.3 | NLP | 0.385 | 0.714 | 0.500 | 0.952 | 1.0 |
|  |  | ICD | **0.462** | **0.600** | **0.522** | **0.947** |  |

## Discussion

Large language models (LLMs) hold immense potential to improve the administration and practice of medicine. In this study, we report the first evaluation of a large language model for zero-shot phenotyping using clinical notes. As a proof-of-concept, we focused on postpartum hemorrhage, the leading cause of maternal morbidity and mortality. We developed a set of 24 postpartum-hemorrhage-related concepts, which included binary identification of a concept or value extraction of a specific measurement. We found that the Flan-T5 model extracted most concepts with high precision and recall, enabling the creation of an interpretable postpartum hemorrhage phenotype that identified significantly more deliveries with postpartum hemorrhage than would be identified by using diagnosis codes alone. Furthermore, these granular concepts can be used for the precise and interpretable identification of the subtypes of postpartum hemorrhage based on the underlying etiology.

Interestingly, we found that the Flan-T5 model can achieve strong performance despite not being specifically trained on specialized EHR data or medical information. This is consistent with prior work that has investigated the utility of GPT-3 in performing few-shot extraction from clinical text [27]. The Flan-T5 performance was best for concrete concepts where the prompt could sufficiently describe how the concept might be described in a clinical note. The model was resilient to misspellings and medical abbreviations. Flan-T5 provided the greatest benefit over regular expressions on more complex concepts, such as coagulation disorders and retained products of conception, which vary in how they are documented. However, the model had lower performance when differentiating closely related concepts, such as manual extraction of the placenta and dilation and curettage, suggesting that clinical instruction-tuning or the use of few-shot exemplars may be beneficial to improve performance.

Our proposed approach demonstrates how complex large language models can be leveraged to create downstream interpretable models. The “extract-then-phenotype” approach enables easy validation of extracted concepts and rapid updating of phenotype definitions. In this work, we chose a postpartum hemorrhage definition that was valid at the time of clinical care [28]. Since 2018, the definition of PPH has changed to include only those cases with blood loss of more than 1000 ml, regardless of the type of delivery [24]. The advantage of our approach is that in such cases, the algorithm can quickly be adapted to remain relevant.

The challenges in identifying patients with PPH are well recognized. Currently, most epidemiological studies rely on ICD diagnostic codes to develop the PPH phenotype. While ICD codes are available as structured data in EHR and many large datasets, the studies using ICD codes demonstrate an underestimation of the incidence of PPH compared to EBL [7] or manual chart review [29]. In addition, studies have demonstrated positive predictive power of 74.5% and only 9% overall prevalence of PPH ICD codes in all patients resenting for cesarean delivery [7]. Zero-shot phenotyping with LLMs enabled us to rapidly identify delivery encounters with postpartum hemorrhage using clinical notes, enabling identification of PPH cases that would have been missed by PPH ICD codes. Our analysis of PPH ICD code prevalence by extracted estimated blood loss (EBL) revealed that there are many clinical notes with EBL values that meet our PPH definition, yet do not have associated PPH ICD codes. There are several possible explanations for this, including preemption of additional blood loss with appropriate medication, inaccurate EBL measurements that do not clinically correlate with the severity of the patient’s presentation, and incorrect extraction of EBL. Nevertheless, these findings underscore the limitations of traditional PPH phenotyping approaches based on claims data alone.

We also created interpretable algorithms to classify different etiology-based subtypes of PPH. This type of analysis is rarely done in other studies as all subtypes are rarely documented [26] Most studies have focused on atonic PPH, which is identified based on the use of uterotonic medications, sometimes available as structured data [30]. In this work, drug administration was not part of the historical medical record and could not be easily retrieved. Our approach using granular concepts from clinical notes allowed for improved classification based on the presence of inadequate uterine tone even when specific medications were not mentioned. Furthermore, this approach is easily portable across institutions as evidenced by the stable performance across two different hospitals in our cohort. The NLP model outperformed an ICD-based approach for three of the four subtypes, only underperforming on the “thrombin” subtype, which had a substantially lower than the other subtypes prevalence of 6.3% in our evaluation cohort. These approaches for subtyping PPH create new opportunities for investigation of different PPH etiologies and for prompt identification of PPH subtypes at the bedside to initiate personalized clinical interventions.

Our work has some limitations. By focusing on discharge notes during the delivery admission, we may have missed cases of delayed or recurrent postpartum hemorrhage, which may have happened later in the postpartum period. Future work could modify the pipeline to analyze later postpartum events. The discharge notes may also reflect institution-specific clinical practices, e.g., the notes may not include some relevant elements such as estimated blood loss and the clinician’s subjective decision to administer medication or perform a procedure. To mitigate this, we selected a long study period and analyzed notes from several hospitals. Another limitation of using Flan-T5 for phenotyping is the size of the model, which, while significantly smaller than many other LLMs, still requires substantial computational resources. Future work could train a clinical instruction-tuned model, which we expect can achieve comparable performance with fewer parameters [31]. Finally, we did not train Flan-T5 on our phenotyping task or consider recent LLMs such as Chat-GPT and GPT-4 in this work. While these models have demonstrated outstanding performance, they are closed models that can require substantial costs when run over lengthy clinical notes. We opt for evaluation of an open-sourced language model which limits security, privacy, and cost concerns.

We investigated the utility of an open-source large language model in performing interpretable, zero-shot clinical phenotyping and subtyping. This approach offers a major advantage compared to existing methods for phenotyping from clinical notes, which rely on manually annotated training data. Zero-shot phenotyping approaches have the potential to accelerate research and improve clinical operations and medicine by enabling the rapid identification of patient cohorts, which are critical for precise disease investigations and the development of personalized medicine tools.

## Methods

### Cohort construction

Mass General Brigham (MGB) is an integrated healthcare system of tertiary and community hospitals and associated outpatient practices across New England. We leveraged MGB’s Research Patient Data Repository (RPDR) [32] to retrieve the data for all MGB patients with a pregnancy related ICD (International Classification of Diseases) or DRG (Diagnoses Related Groups) code according to the enhanced delivery identification method [33]. The available data includes self-reported race and ethnicity, date of birth, insurance information, ICD and DRG codes, and EHR notes. We restricted the cohort to patients with delivery prior to 5/1/2015, when MGB transitioned to using EPIC’s electronic health record system, to enrich for delivery notes with longer narrative sections and less structured data collection. The discharge summaries were filtered to remove duplicates, draft notes, and any notes from the emergency department. The patient’s race, ethnicity, delivery hospital, and age at delivery were extracted from Mass General Brigham’s RPDR. This study was approved by the Mass General Brigham Institutional Review Board, protocol #2020P002859, with a waiver of patient consent.

### Identification of delivery discharge summaries

The discharge summaries retrieved from RPDR could pertain to any hospital admission during the time window. We compared three approaches for identifying the subset of discharge summaries that described an obstetric encounter: a regex approach, an ICD code approach, and a language modelling approach.

The first approach involved the development of a regular expression algorithm. We selected all discharge summaries containing any of the following terms: “labor “, “delivery”, “l&d”, “cesarean section”, “c-section”, “estimated edc”, or “pregnancy”. This approach was designed to optimize for recall, but it may produce false positive examples when there are discharge notes for admissions in the weeks prior and following a delivery. In the second approach, we retrieved all discharge summaries that had a pregnancy-related ICD or DRG code (and do not have an excluded code) within *x* days of the date of the discharge summary, where *x* may be 0, 2, 7, or 14 days. This time-based linking is necessary because there is no encounter linking between billing data and clinical notes in our data repository. In the final approach, we split each note into chunks containing 512 tokens and prompted Flan-T5 with the following question: “Answer the following yes/no question. Does this discharge summary note mention a woman’s delivery? note: note”, where note corresponds to the text of each chunk. If the model responded “yes” for any of the discharge summary sections, we labeled the discharge summary as related to a delivery encounter. See below methods for additional details about the Flan-T5 model

We evaluated each of the approaches on 100 randomly sampled discharge summaries. We found that the language modelling approach yielded the highest overall performance with a macro F1 of 0.936 (Supplemental Table 4) but opted to use the regex approach for creation of the final cohort given its 100% recall.

### Zero-Shot Natural Language Processing to Extract PPH Concepts

We utilized Flan-T5, an instruction-tuned LLM that can be applied to unseen tasks without any additional model training [20], to extract PPH-related concepts from obstetric discharge summaries. Flan-T5 takes as input a prompt describing the task and generates a text output. We leverage Flan-T5 for two broad classes of tasks: binary classification and text extraction. Identification of estimated blood loss is framed as a text extraction task, and all other PPH-related concepts are framed as binary classification tasks.

We developed and evaluated the Flan-T5 models on a set of 1225 manually labelled discharge summaries with ICD codes related to PPH, as defined in Zheutlin et al [8]. We performed stratified sampling to set aside 50 discharge summaries for a small development cohort to conduct minimal prompt engineering for each PPH-related concept, and the remaining 1175 discharge summaries served as our test set. Importantly, there was no overlap in patients between the development and test sets. Furthermore, we only included discharge summaries from a single MGB hospital (Brigham and Women’s Hospital) in the development set in order to evaluate the generalizability of the models to notes from other hospitals in the MGB system.

We performed minimal prompt engineering, constructing one to five prompts for each PPH-related concept. We selected the optimal prompt via binary F1 performance on the development set. Example prompts for PPH-related concepts are in Table 4. For the most complex concepts, such as “placenta accreta spectrum”, “PPH due to surgical causes”, “coagulation disorders with increased risk of bleeding”, and “misoprostol (used as uterotonic)”, we chained together several separate prompts to generate more nuanced predictions. If any of the prompts labeled a concept as present, we considered the concept present.

**Table 4:** Example prompts provided to Flan-T5 to perform zero-shot extraction of postpartum hemorrhage (PPH)-related concepts. We include prompts representative of different types of information retrieval (e.g. presence of a condition, presence of a drug, quantitative value extraction, and multi-prompt concept extraction). “ *<*note*>*” refers to the full text of the note. If the entire note can not fit into a single context window of the model, the text is split into chunks, and the prompt is concatenated to each chunk separately and input into the model. The remaining prompts can be found in our accompanying Github repository. PPH, postpartum hemorrhage; EBL, estimated blood loss.

| Concept | Prompt |
| --- | --- |
| Direct mention of PPH | Answer the following yes/no question. Does the following discharge summary mention **postpartum hemorrhage (PPH)** during the current delivery?<br>note: <note> |
| Methylergonovine | Answer the following yes/no question. Does the following discharge summary mention the **drug methergine (methylergonovine)**?<br>note: <note> |
| Estimated blood loss | Extract the **estimated blood loss (EBL)** in the following discharge summary. If you can't find the answer, please respond "unanswerable".<br>note: <note> |
| Placenta accreta spectrum | Answer the following yes/no question. Does the following discharge summary mention **placenta percreta** during the current delivery?<br>note: <note><br>Answer the following yes/no question. Does the following discharge summary mention **placenta increta** during the current delivery?<br>note: <note><br>Answer the following yes/no question. Does the following discharge summary mention **placenta accreta** during the current delivery?<br>note: <note> |

The length of discharge summaries often exceeded the max length of the input for Flan-T5 models. To address this, we split each discharge summary into text chunks of 512 tokens with a stride of 128, generated a prediction for each text chunk, and aggregated the resulting predictions. For binary classification tasks, we considered the label as positive if any of the text chunks have a positive label. For the text extraction tasks, we took the union of all segments extracted from each text chunk. We also performed post-processing to extract numbers from the generated text. This post-processing was necessary for text extraction tasks because the model can extract random text from the text chunk if a particular text chunk does not contain the information to extract (e.g., a measurement for estimated blood loss).

We experimented with Flan-T5 models of varying sizes on our development set and found that the 11 billion parameter Flan-T5-XXL model performed substantially better than the smaller models (Supplemental Table 5). All analyses in this paper use the Flan-T5-XXL model downloaded from the HuggingFace model hub at https://huggingface.co/google/flan-t5-xxl. Text was generated using greedy decoding with max new tokens set to 5 and temperature of 1.0. We performed inference using a single 16 GB RTX A4000. The Hugging Face accelerate library with DeepSpeed integration was used to speed up inference by enabling a batch size of 48 on a single GPU [34].

We performed inference using Flan-T5 on all delivery-related discharge summaries for each PPH-related concept. We compared the Flan-T5 model to regular expressions constructed for each concept. Regular expressions are the most pertinent baseline because they similarly do not require model training. We compared model performance on the distinct test set of 1175 manually annotated discharge summaries.

### Creation of an NLP-based PPH phenotyping algorithm

During the study period, the hospitals used a PPH definition based on the estimated blood loss (EBL) *>* 500mL during a vaginal delivery or *>* 1000mL during a cesarean delivery [28], and we leveraged the extracted PPH-related concepts to identify PPH deliveries. We focused on the phenotyping of primary PPH, as patients with secondary PPH may develop PPH after discharge [28]. Concretely, we identify a PPH delivery if the sum of all extracted EBL values is *>* 500 and the model predicts a vaginal delivery or if the model sum of all extracted EBL values is *>* 1000 and the model predicts a cesarean delivery. In cases where multiple EBL were extracted with the same value, we assumed that these were multiple mentions of the same EBL in the note and did not sum the values. In cases where the extracted EBL was a single digit, we assumed the EBL was reported in liters and converted the extracted values to milliliters.

We identified PPH deliveries from our entire cohort of delivery discharge summaries and report characteristics of the identified PPH deliveries. We evaluated the positive predictive value of our approach by manually annotating the EBL and cesarean status of a random sample of 300 discharge summaries that the model predicts as describing a PPH delivery. We measured the ability of the NLP-approach to identify PPH cases missed by ICD-based definitions of PPH [7, 8, 35] by calculating the fraction of confirmed PPH cases that have PPH-related ICD codes.

### Creation of an NLP-based PPH subtyping algorithm

We used the extracted PPH-related concepts to develop four composite PPH subtyping algorithms corresponding to distinct causes of PPH: uterine atony (“tone”), retained products of conception, (“tissue”), trauma (“trauma”), and coagulation abnormalities (“thrombin”) [36] (Table 5). We labeled a PPH delivery with the “tone” subtype if the discharge summary contained any of the following: mention of uterine atony, administration of methylergonovine, administration of carboprost or administration of misoprostol as a uterotonic. We labeled a PPH delivery with the “tissue” subtype if the discharge summary contained any of the following: placenta accreta spectrum or retained products of conception, dilatation and curettage, or manual extraction of the placenta. We labeled a PPH delivery with the “trauma” subtype if the discharge summary contained documentation of any of the following: uterine rupture, O’Leary stitches, PPH due to surgical causes (e.g. damage to the uterine artery), or laceration with no other PPH subtype mentioned. We labeled a PPH delivery with the “thrombin” subtype if the discharge summary contained any of the following: mention of disseminated intravascular coagulation, transfusion of platelets, transfusion of cryoprecipitate, or transfusion of frozen fresh plasma in ratio with red blood cells higher than 1:1:1.

**Table 5:** Postpartum hemorrhage (PPH) subtype definitions. The natural language processing (NLP)-based approach that leverages the extracted PPH concepts is on the left. A note is labelled with a given subtype if any of the concepts are present in the note. One exception is the “laceration” concept, which only signifies the “trauma” subtype if no other subtypes have been assigned to the note. The baseline international classification of diseases (ICD)-based approach is on the right. Similarly, a note is assigned to a subtype if any of the ICD codes are present. Here *x* refers to any number 0 *−* 9, meaning that any more specific ICD code under that in the ICD ontology will suffice. For example, 666.14 is a valid ICD code for the “tone“ subtype. 664.x only signifies the “trauma” subtype if no 666.x ICD code is present (*i.e.* there are no other attributable causes of the hemorrhage). These rules for the “laceration” subtype are designed to mitigate false positives due to lacerations during the delivery that do not result in significant hemorrhage. NLP, natural language processing; PPH, postpartum hemorrhage; ICD, international classification of diseases.

| Subtype | NLP-based definition | ICD-based definition |
| --- | --- | --- |
| Tone | uterine atony; methergine / methylergonovine; carboprost; misoprostol as a uterotonic | 666.1x |
| Tissue | placenta accreta spectrum; retained products of conception; dilatation and curettage; manual extraction of the placenta | 666.0x |
| Trauma | uterine rupture; PPH due to surgical causes; O’Leary stitches; laceration (if no other subtype is valid) | 665.x or (664.x if no 666.x) |
| Thrombin | coagulation disorders with increased risk of bleeding; administration of platelets, fresh frozen plasma, or cryoprecipitate in a ratio with red blood cells higher than 1:1:1 | 666.3x |

We performed subtyping on all deliveries that the NLP-based model predicted as describing postpartum hemorrhage. We validated the performance of the classification algorithms via manual chart review of the confirmed PPH deliveries identified above (See “Identification of deliveries with PPH”). We compared the NLP-based approach to a subtyping approach that leveraged ICD codes. The ICD definitions for each subtype based on conversion from ICD10 codes [26] are in Table 5.

### Validation of model performance

We performed manual annotation of discharge summaries to assess the performance of our models for extracting PPH-related concepts, identifying patients with PPH, and classifying subtypes of PPH. In each use case, one of four annotators with expe-rience in reading obstetrics notes (M.R., R.F., A.G., and V.K.) first annotated the notes with the relevant PPH-related concepts or subtypes, and a clinician (V.K. and K.G.) subsequently reviewed all annotations.

We extended PRAnCER (Platform Enabling Rapid Annotation for Clinical Entity Recogni-tion) [37], an annotation tool for clinical text, to collect annotations for the PPH-related concepts. During the annotation process, the annotator highlights a span of text in the note and assigns it to the corresponding label. Regular expressions are used initially to highlight potential annota-tions, and the annotator can confirm or reject the regex “pre-annotation”. We found that this pre-annotation process can accelerate the annotation workflow and improve annotator recall. However, it does have the potential to induce over-reliance on the pre-annotations. An image of the modified annotation tool is found in Supplemental Figure 1.

### Statistical analyses

We performed a McNemar test to assess whether there is a significant dif-ference in performance between our NLP-based approach and a regular expression approach for extracting PPH-related concepts. We similarly used a McNemar test to compare the NLP-based and ICD-based approaches for subtyping PPH deliveries.

## Data availability

The MGB data used in this study contains identifiable protected health in-formation and therefore cannot be shared publicly. MGB investigators with an appropriate IRB approval can contact the authors directly regarding data access.

## Data Availability

The patient data used in this study contains identifiable protected health information and therefore cannot be shared publicly. Mass General Brigham investigators with an appropriate IRB approval can contact the authors directly regarding data access.

## Acknowledgements

KJG reports funding from NIH/NHLBI grants K08 HL146963-02, K08 HL146963-02S1, and R03 HL162756. VPK reports funding from the NIH/NHLBI grants 1K08HL161326-01A1, Anesthesia Patient Safety Foundation (APSF), and BWH IGNITE Award. The funders played no role in study design, data collection, analysis and interpretation of data, or the writing of this manuscript.

