## Supplemental Figures and Tables for "Zero-shot Interpretable Phenotyping of Postpartum Hemorrhage Using Large Language Models"

| Concept Type | Concept | Annotation instructions |
|---|---|---|
| Medication | Carboprost | carboprost (hemabate) given for inadequate uterine tone, example 'hemabate' |
| | Methylergonovine | methylergonovine (methergine) given for inadequate uterine tone, example 'methergine' |
| | Misoprostol as a utero-tonic | misoprostol (cytotec) given for inadequate uterine tone after delivery; do not annotate when given to induce labor (in this case, it is administered before delivery); example 'cytotec' |
| Procedure | Bakri balloon | Bakri balloon or hemostatic balloon inserted in the uterus to stop bleeding, example 'Bakri balloon' |
| | Dilation and curettage | procedure performed after delivery to remove retained products of conception; examples: 'Dilation & curettage', 'D&C', 'D&E', 'Dilation and evacuation', 'D and E'. |
| | Manual extraction of placenta | manual extraction/evacuation/removal of the placenta or retained products of conception |
| | O'Leary sutures | O'Leary sutures performed on the uterus to stop bleeding, example 'O'Leary' |
| | Hysterectomy | hysterectomy, or surgical resection of the uterus; examples 'C-hyst', 'Cesarean hysterectomy', 'hyst'. |
| Transfusion | Fresh frozen plasma | transfusion of fresh frozen plasma, examples 'ffp', 'FFP', etc. |
| | Cryoprecipitate | transfusion of cryoprecipitate, examples 'cryo', 'CP', etc. |
| | Platelets | transfusion of platelets, examples 'plt', 'Pt', etc. |
| | Packed red blood cells | transfusion of packed red blood cells, examples 'prbc', 'rbc', etc. |
| Problem | Uterine atony | inadequate uterine tone; examples 'atony', 'poor uterine tone' |
| | Laceration | laceration or tear, example 'lac' |
| | Uterine rupture | uterine rupture or uterine dehiscence, example 'uterine rupture' |
| | Retained products of conception | retained products of conception, retained placenta, or retained tissue, example 'rPOC' |

| | | |
|---|---|---|
| | Placenta accreta spectrum | an abnormally adherent placenta, or placenta accreta, percreta, or increta, example 'accreta' |
| | Abruption of the placenta | abruption or separation of the placenta before delivery, example 'abruption' |
| | Placenta previa | placenta previa, include 'partial previa' but only if present at delivery, do not include if resolved, example 'previa' |
| | Coagulation disorders | Coagulation disorders associated with a higher risk of bleeding, such as thrombocytopenia, HELLP, coagulation factor deficiency, and disseminated intravascular coagulopathy, which can be represented as 'DIC', 'coagulopathy' or 'bleeding/oozing from all sites' |
| Problem | PPH due to surgical causes | PPH due to surgical trauma during delivery; examples, 'an extension of the uterine incision', 'laceration of uterine artery' |
| | Direct mention of PPH | Explicit documentation of postpartum hemorrhage or hemorrhage after delivery, example 'pph' |
| Delivery type | Cesarean delivery | Cesarean mode of delivery, example 'c-section' |

**Table S1: List of postpartum hemorrhage (PPH)-related concepts that are extracted from obstetric discharge summaries and the descriptions for each concept provided to annotators.** PPH, postpartum hemorrhage.

| Concept (n) | Sensitivity | Specificity | PPV | Binary F1 | Acc. |
|---|---|---|---|---|---|
| Carboprost (n=49) | 0.980 | 1.000 | 1.000 | 0.990 | 0.999 |
| Laceration (n=421) | 0.971 | 0.987 | 0.976 | 0.974 | 0.981 |
| Bakri balloon (n=46) | 0.957 | 0.999 | 0.978 | 0.967 | 0.997 |
| Fresh frozen plasma (n=79) | 1.000 | 0.994 | 0.919 | 0.958 | 0.994 |
| Uterine atony (n=107) | 0.888 | 0.999 | 0.990 | 0.936 | 0.989 |
| Cesarean delivery (n=470) | 0.915 | 0.965 | 0.945 | 0.930 | 0.945 |
| Cryoprecipitate (n=29) | 1.000 | 0.996 | 0.853 | 0.921 | 0.996 |
| Methylergonovine (n=104) | 0.865 | 0.998 | 0.978 | 0.918 | 0.986 |
| Hysterectomy (n=55) | 1.000 | 0.991 | 0.846 | 0.917 | 0.991 |
| Placenta previa (n=170) | 0.900 | 0.989 | 0.933 | 0.916 | 0.976 |
| Misoprostol as a uterotonic (n=112) | 0.982 | 0.972 | 0.786 | 0.873 | 0.973 |
| O'Leary sutures (n=12) | 0.833 | 0.999 | 0.909 | 0.870 | 0.997 |
| Packed red blood cells (n=165) | 0.909 | 0.965 | 0.811 | 0.857 | 0.957 |
| Uterine rupture (n=5) | 1.000 | 0.998 | 0.714 | 0.833 | 0.998 |
| Direct mention of PPH (n=362) | 0.732 | 0.986 | 0.960 | 0.831 | 0.908 |
| Retained products of conception (n=102) | 0.647 | 0.991 | 0.868 | 0.742 | 0.961 |
| Placenta accreta spectrum (n=45) | 0.667 | 0.992 | 0.769 | 0.714 | 0.980 |
| Dilation and curettage (n=122) | 0.648 | 0.971 | 0.718 | 0.681 | 0.937 |
| Platelets (n=29) | 0.862 | 0.983 | 0.556 | 0.676 | 0.980 |
| PPH due to surgical causes (n=27) | 0.407 | 0.999 | 0.917 | 0.564 | 0.986 |
| Coagulation disorders (n=44) | 0.364 | 0.997 | 0.842 | 0.508 | 0.974 |
| Abruption of the placenta (n=47) | 0.340 | 0.996 | 0.800 | 0.478 | 0.970 |
| Manual extraction of placenta (n=110) | 0.309 | 0.999 | 0.971 | 0.469 | 0.934 |
| **Concept (n)** | **Sensitivity** | **PPV** | **Note Acc.** | | |
| Estimated blood loss (n=212) | 0.535 | 0.737 | 0.884 | | |

**Table S2: Performance of regular expressions assessed on 1,075 manually annotated discharge summaries.** PPH, postpartum hemorrhage.

| **Concept** | **Hospital** | **Count** | **Binary F1** |
|---|---|---|---|
| PPH | BWH | 195 | 0.891 |
| | MGH | 164 | 0.903 |
| PPH due to surgical causes | BWH | 11 | 0.381 |
| | MGH | 16 | 0.711 |
| Placental abruption | BWH | 38 | 0.872 |
| | MGH | 8 | 0.941 |
| Cesarean section | BWH | 331 | 0.963 |
| | MGH | 132 | 0.985 |
| Misoprostol as a uterotonic | BWH | 94 | 0.916 |
| | MGH | 17 | 0.971 |
| Dilation and curettage | BWH | 91 | 0.812 |
| | MGH | 29 | 0.846 |
| Coagulation Disorders | BWH | 35 | 0.870 |
| | MGH | 8 | 0.769 |
| Fresh frozen plasma | BWH | 74 | 0.953 |
| | MGH | 5 | 0.909 |
| Carboprost | BWH | 33 | 0.970 |
| | MGH | 15 | 0.846 |
| Hysterectomy | BWH | 42 | 0.955 |
| | MGH | 12 | 0.815 |
| Laceration | BWH | 258 | 0.984 |
| | MGH | 163 | 0.985 |
| Manual extraction of placenta | BWH | 86 | 0.740 |
| | MGH | 23 | 0.824 |
| Methylergonovine | BWH | 64 | 0.953 |
| | MGH | 39 | 0.974 |
| Packed red blood cells | BWH | 128 | 0.907 |
| | MGH | 37 | 0.850 |
| Placenta accreta spectrum | BWH | 37 | 0.846 |
| | MGH | 8 | 0.421 |
| Placenta previa | BWH | 121 | 0.940 |

| | | | |
|---|---|---|---|
| | MGH | 47 | 0.979 |
| Retained products of conception | BWH | 68 | 0.897 |
| | MGH | 32 | 0.933 |

**Table S3:** Comparison of Flan-T5 Large Language Model (LLM) performance in extracting postpartum hemorrhage (PPH)-related concepts across Mass General Brigham hospital sites. All prompt engineering was performed exclusively on notes from Brigham and Women's Hospital, and the remaining hospitals serve as external test sets. Sites with at least 5 notes with the concept are included, and only labels with more than one site are shown. PPH, postpartum hemorrhage; BWH, Brigham and Women's Hospital, MGH, Massachusetts General Hospital.

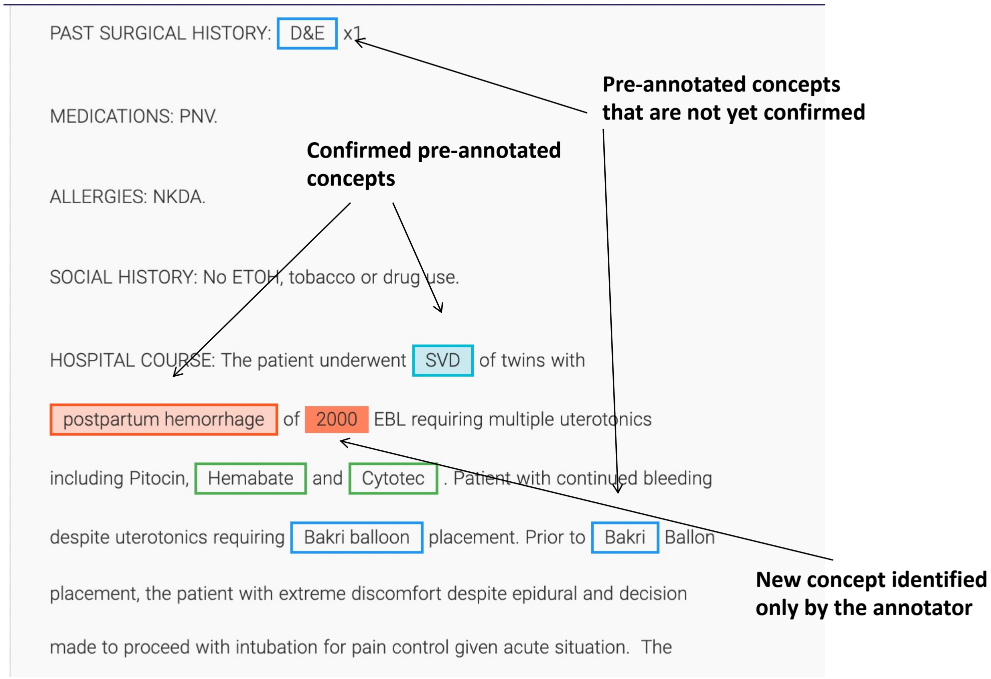


**Figure S1: An image capture of the modified PRAnCER annotation tool.**

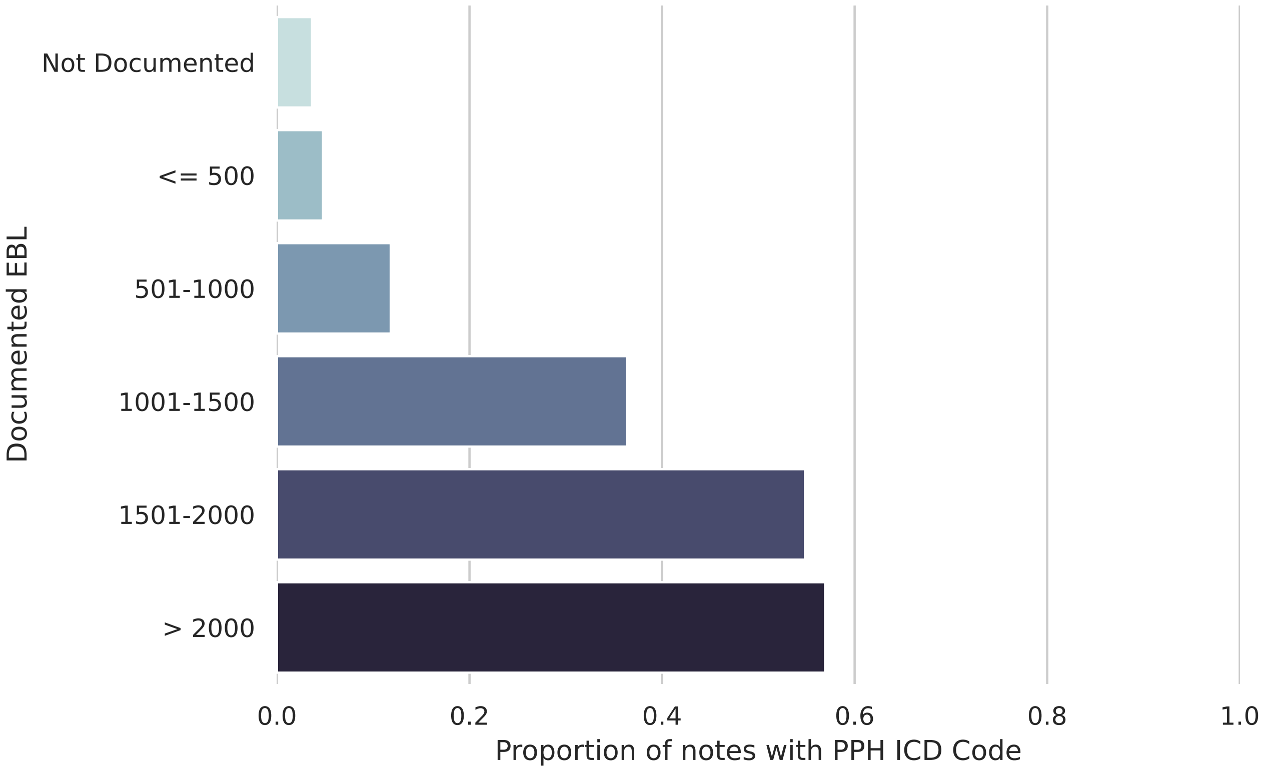


**Figure S2: Prevalence of postpartum hemorrhage( PPH) international classification of diseases (ICD) codes in notes with varying estimated blood loss values documented.** The $y$-axis depicts the range of estimated blood loss (EBL) values extracted from the delivery note by Flan-T5 (if any), and the $x$-axis denotes the proportion of notes in each EBL category with a postpartum hemorrhage ICD diagnostic code. PPH ICD codes are defined according to the definition in Butwick et al. Refer to Figure 3 for a similar plot using the PPH ICD definition in Zheutlin et al. PPH, postpartum hemorrhage; ICD, international classification of diseases; EBL, estimated blood loss.

| Model | Sensitivity | Specificity | PPV | Binary F1 | Acc. |
|---|---|---|---|---|---|
| Flan-T5 Zero Shot | 0.986 | 0.857 | 0.947 | | **0.950** |
| Regex | **1.000** | 0.643 | 0.878 | | 0.900 |
| ICD Code - Day of Note | 0.472 | **1.000** | **1.000** | | 0.620 |
| ICD Code - Within 2 Days | 0.847 | 0.964 | 0.984 | | 0.880 |
| ICD Code - Within 7 Days | 0.944 | 0.857 | 0.944 | | 0.920 |
| ICD Code - Within 14 Days | 0.972 | 0.857 | 0.946 | | 0.940 |

**Table S4:** Comparison of approaches for identifying discharge notes that describe a labor and delivery encounter on a random sample of 100 discharge summaries. ICD, international classification of diseases.

| Model Size | Sensitivity | Specificity | PPV | Binary F1 | Acc. |
|---|---|---|---|---|---|
| Small | 0.833 | 0.136 | 0.116 | 0.204 | 0.220 |
| Base | 0.500 | 0.568 | 0.136 | 0.214 | 0.560 |
| Large | 0.333 | **0.977** | 0.667 | 0.444 | 0.900 |
| XL | **1.000** | 0.773 | 0.375 | 0.545 | 0.800 |
| XXL | **1.000** | 0.955 | **0.750** | **0.857** | **0.960** |

**Table S5:** Impact of Flan T5 model size on zero shot classification performance for identifying mentions of postpartum hemorrhage in notes. All metrics are reported on the small prompt tuning set of 50 notes.
